# Post-Vaccination Symptoms after A Third Dose of mRNA SARS-CoV-2 Vaccination in Patients with Inflammatory Bowel Disease

**DOI:** 10.1101/2021.12.05.21266089

**Authors:** Dalin Li, Philip Debbas, Susan Cheng, Jonathan Braun, Dermot P.B. McGovern, Gil Y. Melmed

**Affiliations:** Inflammatory Bowel and Immunobiology Research Institute, Karsh Division of Digestive and Liver Diseases, Department of Medicine, Cedars-Sinai Medical Center, Los Angeles, CA; Smidt Heart Institute, Department of Medicine, Cedars-Sinai, Los Angeles, CA; Samual Oschin Comprehensive Cancer Institute, Cedars-Sinai, Los Angeles, CA; Children’s Hospital, Orange County, CA; Banner University Medical Center, Phoenix, AZ; University of California, San Diego, CA; Sibley Memorial Hospital, Johns Hopkins, Washington, DC; Swedish Medical Center, Seattle, WA; Capital Digestive Care, Chevy Chase, MD; University of Utah, Salt Lake City, UT; Temple University, Philadelphia, PA; UT Southwestern, Dallas, TX; Center for Rheumatology, Los Angeles, CA; Baylor College of Medicine, Houston, TX; Hoag Hospital, Newport Beach, CA; Johns Hopkins Hospital, Baltimore, MD; The Oregon Clinic, Portland, OR; Sutter Health, Palo Alto, CA; Medstar-Georgetown, Washington, DC; Saratoga-Schenectady Gastroenterology, Saratoga Springs, NY; University of California, Irvine, CA; The Mayo Clinic, Rochester, MN; University of Chicago, Chicago, IL; Keck Medicine of University of Southern California, Los Angeles, CA; Dartmouth-Hitchcock Medical Center, Lebanon, NH; Atlanta Gastroenterology Associates, Atlanta, GA; Gastro One, Germantown, TN; University of Miami, Miami, FL

## Abstract

Symptoms after SARS-CoV-2 primary vaccination among patients with inflammatory bowel disease (IBD) are generally of similar frequency, severity, and duration to those reported in the general population. The symptom profile after a 3^rd^ mRNA vaccine dose in the predominantly immune-compromised IBD population is unknown. We aimed to assess symptomology after a 3^rd^ or booster dose of mRNA vaccination in adults with IBD. We surveyed participants of the Coronavirus Risk Associations and Longitudinal Evaluation in IBD (CORALE-IBD) post-vaccination registry for symptom frequency and severity after a 3^rd^ mRNA vaccine dose in an observational cohort study. In total, 524 participants (70% female, mean age 45 years) reported a third dose of mRNA vaccination through October 11, 2021. Overall, 41% reported symptoms after a third dose, with symptoms generally more frequent and more severe among participants younger than 55 years. The most frequent postvaccination symptoms were injection site pain (39%), fatigue or malaise (34%), and headache (23%). These symptoms were all less frequently reported after dose 3 than after dose 2. Gastrointestinal symptoms were reported by 8.8%, which was slightly more frequent than after dose 2 (7.8%). Those with severe symptoms after dose 2 were more likely to have severe symptoms after dose 3. These findings can reassure the IBD patient and provider communities that the likelihood and distribution of symptoms after a third mRNA vaccine dose are generally similar to those after a second dose, and that the frequency of postvaccination symptoms after dose 3 are generally lower than after dose 2.

## Introduction

Vaccine safety concerns are a major contributor to vaccine hesitancy.^1^ Symptoms after SARS-CoV-2 primary vaccination among patients with inflammatory bowel disease (IBD) are generally of similar frequency, severity, and duration to those reported in the general population,^2, 3^ although adverse events after the second dose of a two-dose primary inoculation are more frequent and severe than after the first dose.^4^ In the general population, the frequency of reactions after a third dose of mRNA vaccination in the general population was similar to the second dose.^5^ However, the symptom profile after a 3^rd^ mRNA vaccine dose in the predominantly immune-compromised IBD population is unknown. We aimed to assess symptomology after a 3^rd^ or booster dose of mRNA vaccination in adults with IBD.

## Methods

Adults with IBD participating in the prospective Coronavirus Risk Associations and Longitudinal Evaluation in IBD (CORALE-IBD) vaccine registry were queried after FDA approval of a 3^rd^ dose of mRNA vaccine for patients receiving immune suppressive therapy. Participants who received a 3^rd^ dose were asked to complete a detailed survey about post-vaccination symptoms 1 week after vaccination. Given that symptoms after the second dose of the primary series were more frequent and severe than after the first dose, we compared the frequency and severity of symptoms after dose 3 relative to those reported after dose 2 (R version 3.5.1). Symptoms were assessed across eleven organ systems, including injection site symptoms (pain, erythema, swelling); fatigue or malaise; headache, dizziness or lightheadedness; fever or chills; rheumatologic (muscle, joint, or nerve) symptoms, gastrointestinal (nausea, vomiting, abdominal pain, diarrhea, or other) symptoms; sleep changes; swollen lymph nodes; skin/nail or facial changes; eye, ear, mouth or throat changes; cough, chest or breathing symptoms, and memory or mood changes. Symptoms were graded by severity as mild, moderate, or severe impact on activities of daily living, or requiring hospitalization as defined in Baden et al^3^. The category of “severe+” included those with severe symptoms, as well as those with need for hospitalization. Given the distinct symptom profiles experienced by older people relative to younger vaccine recipients after dose 2, we stratified patients by age less than or ≥50. We also constructed a 3×3 severity table (none-mild, moderate, and severe+) to clarify the frequency of severe symptoms after dose 3 relative to the severity of symptoms experienced after dose 2. The study protocol was approved by the Cedars-Sinai institutional review board. All study participants provided informed consent.

## Results

The cohort included 524 participants (70% female, mean age 45 years) reporting a third dose of mRNA vaccination through October 11, 2021. A majority had Crohn’s disease (71%) with the remainder having ulcerative colitis or IBD-unclassified, and 89% were receiving biologic therapies. Most participants (58%) received primary vaccination with BNT562b2, and only 3.5% of the overall cohort reported a previous COVID infection at the time of initial vaccination. Overall, 97% of subjects received a third dose with the same mRNA vaccine as in their initial series with the remainder receiving the other mRNA vaccine type. No participants received a third dose with the Ad26.CoV.2 (J&J) vaccine.

Overall, 41% reported symptoms after a third dose, with symptoms generally more frequent and more severe among participants younger than 55 years (**Supplemental Table 1**). The most frequent postvaccination symptom was injection site pain (39%). Commonly reported systemic symptoms were fatigue or malaise (34%), headache (23%), and muscle, bone and joint symptoms (13%). These symptoms were all less frequently reported after dose 3 than after dose 2 (**Figure**). Gastrointestinal symptoms were reported by 8.8%, which was slightly more frequent than after dose 2 (7.8%). Among those with postvaccination symptoms, the proportion with severe+ symptoms after dose 3 was lower than dose 2 for fatigue/malaise, headache, dizziness and lightheadedness, fever or chills, and rheumatologic symptoms, but was slightly higher than dose 2 for gastrointestinal symptoms (**Figure**).

**Figure:**
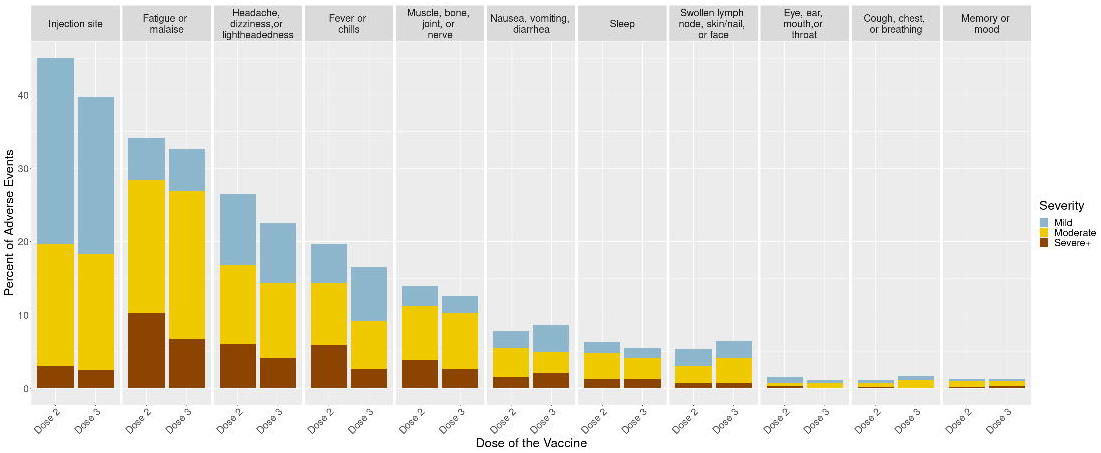
Severity of Symptoms by System for Dose 3 relative to Dose 2

We evaluated the severity of symptoms after dose 3 by the severity of symptoms after dose 2 (**Supplemental Table 2**). Overall, the majority (75%) experienced none or mild symptom severity for both doses. Severe+ symptoms were comparable at dose 2 and 3 (17% and 14%, respectively). Of those with severe+ symptoms after dose 2, 34% experienced severe+ symptoms after dose 3 (OR 5.15, p<0.001). In comparison, about 22% experienced severe+ symptoms after dose 3 but did not report severe+ symptoms after dose 2.

## Discussion

We demonstrate several key findings with respect to symptoms after a third mRNA vaccine among patients with inflammatory bowel disease. First, post-vaccination symptom frequency and severity are significantly greater among those younger than 55 years. This is similar to findings after a second vaccine dose. Second, symptoms after dose 3 were slightly less frequent and less severe for most organ systems, with the notable exception of gastrointestinal symptoms which were slightly more common and severe after dose 3 relative to dose 2. Third, the frequency of severe+ symptoms were comparable after dose 2 and dose 3. They occurred in one third of those who experienced them after dose 2, and in about 22% of those who did not experience them after dose 2. This suggests that most people with severe+ symptoms after dose 2 do not get severe symptoms after a 3^rd^ dose, but still have a higher likelihood of having severe+ symptoms than those who did not have severe+ symptoms after dose 2. Because about one in five experience severe+ symptoms after dose 3 even without previous severe+ symptoms, patients should consider vaccination at a time when short-lived severe symptoms can be best tolerated and addressed.

Overall, our findings can reassure the IBD patient and provider communities that the likelihood and distribution of symptoms after a third mRNA vaccine dose are generally similar to those after a second dose, and that for most organ systems, fewer people experienced postvaccination symptoms after dose 3 than after dose 2. Further evaluation of postvaccination gastrointestinal symptoms in this population living with chronic gastrointestinal illness is warranted.

## Data Availability

All data produced in the present study are available upon reasonable request to the authors

## Acknowledgements

We thank all patient participants of the CORALE-IBD Vaccine study.

