## Supplemental Tables for "Post-Vaccination Symptoms after A Third Dose of mRNA SARS-CoV-2 Vaccination in Patients with Inflammatory Bowel Disease"

**Supplemental Table 1:** Frequency and Severity of Postvaccination Symptoms by Age

| variable | Total | Age<55 | Age>=55 | P |
| --- | --- | --- | --- | --- |
| N | 524(100) | 387(100) | 137(100) |  |
| Age | 45.3(14.44) | 38.07(8.24) | 65.72(6.33) | 2.09E-267 |
| Female | 370(70.75) | 293(75.91) | 77(56.2) | 7.17E-05 |
| Crohn's disease | 370(70.61) | 274(70.8) | 96(70.07) | 0.872 |
| White | 491(93.88) | 360(93.26) | 131(95.62) | 0.422 |
| Primary vaccine: BNT162b2 (Pfizer/BioNtech) | 306(58.4) | 231(59.69) | 75(54.74) | 0.313 |
| Prior COVID | 18(3.45) | 15(3.9) | 3(2.19) | 0.347 |
| On biologic therapy at time of vaccination | 467(89.12) | 347(89.66) | 120(87.59) | 0.503 |
| Duration between doses 2 and 3 (days, SD)) | 166.00(32.36) | 164.19(34.11) | 170.80(26.76) | 0.456 |
| Adverse Events |  |  |  |  |
| Local pain | 206(39.3) | 167(43.15) | 39(28.46) | 2.49E-03 |
| Local redness | 58(11.07) | 47(12.14) | 11(8.03) | 0.187 |
| Local swelling | 58(11.07) | 49(12.66) | 9(6.57) | 0.051 |
| Fever or chills | 87(16.6) | 72(18.6) | 15(10.95) | 0.038 |
| Fatigue or malaise | 176(33.59) | 142(36.69) | 34(24.82) | 0.011 |
| Headache | 119(22.71) | 97(25.06) | 22(16.06) | 0.031 |
| Eye, ear, mouth or throat symptoms | 11(2.1) | 11(2.84) | 0(0.00) | 0.046 |
| Lymph node, skin or facial symptoms | 36(6.87) | 31(8.01) | 5(3.65) | 0.083 |
| Cough, chest or breathing symptoms | 9(1.72) | 7(1.81) | 2(1.46) | 0.787 |
| Digestive symptoms | 46(8.78) | 40(10.34) | 6(4.38) | 0.034 |
| Urinary or genital | 0(0.00) | 0(0.00) | 0(0.00) | 1.000 |
| Muscle, bone or joint | 66(12.6) | 54(13.95) | 12(8.76) | 0.115 |
| Memory or mood | 7(1.34) | 6(1.55) | 1(0.73) | 0.472 |
| Sleep symptoms | 31(5.92) | 28(7.24) | 3(2.19) | 0.031 |
| Overall Severity |  |  |  |  |
| None | 307(58.59) | 213(55.04) | 94(68.61) | 0.026 |
| Mild | 30(5.73) | 25(6.46) | 5(3.65) |  |
| Moderate | 121(23.09) | 93(24.03) | 28(20.44) |  |
| Severe+ | 66(12.6) | 56(14.47) | 10(7.3) |  |

**Supplemental Table 2:** Severity of Symptoms after Dose 3 by Severity of Symptoms after Dose 2

|  |  | Dose 2 AE(N/%) | |
| --- | --- | --- | --- |
| Dose 3 AE | None/Mild | Moderate | Severe+ |
| None/Mild | 261(73.31) | 61(51.26) | 41(45.56) |
| Moderate | 68(19.1) | 41(34.45) | 18(20) |
| Severe+ | 27(7.58) | 17(14.29) | 31(34.44) |

AE: Adverse events

Severe+: Severe symptoms, or symptoms requiring hospitalization
